# *In vivo* detection of substantia nigra and locus coeruleus volume loss in Parkinson’s disease using neuromelanin-sensitive MRI: Replication in two cohorts

**DOI:** 10.1101/2022.02.23.22271356

**Authors:** Kristy S. Hwang, Jason Langley, Richa Tripathi, Xiaoping P. Hu, Daniel E. Huddleston

**Author notes:** These authors contributed equally to this work (Kristy S. Hwang, Jason Langley).

## Abstract

Patients with Parkinson’s disease undergo a loss of melanized neurons in substantia nigra pars compacta and locus coeruleus. Very few studies have assessed substantia nigra pars compacta and locus coeruleus pathology in Parkinson’s disease simultaneously with magnetic resonance imaging (MRI). Neuromelanin-sensitive MRI measures of substantia nigra pars compacta and locus coeruleus volume based on explicit magnetization transfer contrast have been shown to have high scan-rescan reproducibility in controls, but no study has replicated detection of Parkinson’s disease-associated volume loss in substantia nigra pars compacta and locus coeruleus in multiple cohorts with the same methodology.

Two separate cohorts of Parkinson’s disease patients and controls were recruited from the Emory Movement Disorders Clinic and scanned on two different MRI scanners. In cohort 1, imaging data from 19 controls and 22 Parkinson’s disease patients were acquired with a Siemens Trio 3 Tesla scanner using a 2D gradient echo sequence with magnetization transfer preparation pulse. Cohort 2 consisted of 33 controls and 39 Parkinson’s disease patients who were scanned on a Siemens Prisma 3 Tesla scanner with a similar imaging protocol. Locus coeruleus and substantia nigra pars compacta volumes were segmented in both cohorts.

Substantia nigra pars compacta volume (Cohort 1: *p*=0.0148; Cohort 2: *p*=0.0011) and locus coeruleus volume (Cohort 1: *p*=0.0412; Cohort 2: *p*=0.0056) were significantly reduced in the Parkinson’s disease group as compared to controls in both cohorts. This imaging approach robustly detects Parkinson’s disease effects on these structures, indicating that it is a promising marker for neurodegenerative neuromelanin loss.

## Introduction

Parkinson’s disease (PD) is a heterogeneous neurodegenerative disorder with a variety of motor and non-motor symptoms that can be clinically challenging to diagnose and manage, and there are currently no effective interventions to stop PD neurodegeneration. Postmortem studies have yielded some insights into PD biology, and by the time of symptom onset and clinical diagnosis, there is an estimated 40-60% loss of pigmented dopamine neurons in the substantia nigra compacta (SNc) [1-3]. Neuromelanin loss in both the SNc and the locus coeruleus (LC) are hallmark pathologies in PD [3-5]. However, the role of neuromelanin in PD pathogenesis has been challenging to study due to a lack of tools to investigate neuromelanin biology in living patients. Studies in rodents suggest that extracellular neuromelanin triggers a neuroinflammatory cascade that accelerates neurodegeneration [6, 7]. Emerging evidence from a new transgenic rodent model that expresses neuromelanin in SNc indicates that neuromelanin dynamics may play a key role in neurodegeneration, and neuromelanin modulating therapies for PD have been proposed [8-10]. Neuroprotection trial designs could be improved with brain imaging markers of neurodegeneration, including markers of change in tissue neuromelanin. These imaging markers might assist participant selection and serve as surrogate outcome measures for neuroprotection trials. Therefore, robust and reproducible neuroimaging measures are needed to detect and quantify PD-associated neuromelanin changes *in vivo*.

Melanized neurons in LC and SNc can be visualized *in vivo* with neuromelanin-sensitive MRI (NM-MRI) sequences using either incidental magnetization transfer effects from an interleaved multi-slice turbo spin echo acquisition [11] or explicit magnetization transfer effects generated by magnetization transfer preparation pulses [12-14]. Magnetization transfer contrast (MTC) colocalizes with melanized neurons in LC and SNc [15, 16]. NM-MRI approaches using incidental [17] or explicit [18-20] magnetization transfer effects have been found to exhibit high scan-rescan reproducibility in controls, and gradient echo-based approaches with explicit magnetization transfer effects exhibit the highest reproducibility [20].

NM-MRI can be used to assess PD-related reductions in tissue neuromelanin content in LC and SNc. Application of NM-MRI approaches based on incidental magnetization transfer effects to image depigmentation has revealed PD-related reductions in NM-MRI contrast ratios in SNc or LC [11, 21, 22], nigral volume [23, 24], or the area of SNc in a single slice [25, 26]. Similar reductions have been observed in nigral volume [12] and contrast using gradient echo NM-MRI sequences that measure explicit magnetization transfer effects [27]. Nigral regions of interest, derived from NM-MRI images, have also been used to examine PD-related microstructural changes [28-30] or iron deposition [31, 32] in SNc.

Replication of imaging markers in multiple cohorts is a crucial step in the development of translationally useful methods. Demonstrating reliable detection of neuromelanin loss in LC and SNc would help better establish NM-MRI measures as informative to the study of PD, particularly for investigation of the role of neuromelanin in its pathogenesis. Candidate markers with established reproducibility would warrant further study in larger longitudinal studies, and they may ultimately be applied in clinical trials for neuromelanin modulating therapies in PD and related conditions. Reliable imaging of LC is particularly important for prodromal detection strategies, because LC may be involved earlier in the disease course than SNc [33].

Reductions in nigral volume have been replicated in separate cohorts using explicit magnetization transfer effect-based NM-MRI approaches [34]. However, that study did not include LC volume measurement, and PD-related reductions in LC volume have not been replicated separately either. This deficiency may be due to the size and stature of LC, a small rod-shaped structure approximately 2 mm in diameter and 15 mm long [35]. Here we aim to remedy this deficiency by applying a NM-MRI approach based on explicit magnetization transfer effects to measure LC and SNc volumes in PD patients and controls in two separate cohorts acquired on different MRI scanner models.

## Methods

### Research Participants

All research participants were recruited from the Emory University Movement Disorders Clinic under an institutional review board approved protocol with informed written consent and in accordance with the Declaration of Helsinki. Two cohorts were studied in this research, and the cohorts were scanned on two different MRI scanners as described below. Cohort 1 data was collected from 2012-2014 and included 19 controls and 22 PD patients. Cohort 2 consisted of 33 healthy controls (HC) and 39 PD patients with data collected from 2015-2016. Controls were recruited from the community and the Emory Alzheimer’s Disease Research Center control population. PD patients fulfilled the Movement Disorders Society clinical diagnostic criteria [36], and diagnosis was established by a fellowship-trained movement disorders neurologist at the Emory Movement Disorders Clinic. Specific exclusion criteria included the following: 1) PD patients showing symptoms or signs of secondary or atypical parkinsonism [37], 2) controls were excluded if they scored ≤26 on the Montreal Cognitive Assessment (MOCA) indicating cognitive impairment, 3) any history of vascular territorial stroke, epilepsy, multiple sclerosis, neurodegenerative disease (aside from PD), peripheral neuropathy with motor deficits, parenchymal brain tumor, hydrocephalus, or schizophrenia, 4) treatment with an antipsychotic drug (other than quetiapine at a dose less than 200mg daily), or 5) if there were any contraindications to MRI imaging.

PD participants had early to moderate disease with Unified Parkinson’s Disease Rating Scale Part III (UPDRS-III) motor score ≤25 in the practically defined ON state. Disease duration (in years) was defined as scan date subtracted from the date of disease onset and levodopa equivalent daily dose (LEDD) was also determined for PD participants. Cognition was assessed using the Montreal Cognitive Assessment (MoCA) [38]. The Non-motor Symptoms Questionnaire (NMSQ) [39] was used to assess non-motor symptoms. Symptoms of rapid eye movement (REM) behavior disorder were assessed using the REM Sleep Behavior Disorder Screening Questionnaire (RBD-SQ) [40]. Participants were evaluated on the same day as the MRI scan.

### MRI Acquisition

MRI data for Cohort 1 were acquired with a Siemens Trio 3 Tesla scanner (Siemens Medical Solutions, Malvern, PA, USA) at Emory University with a 12-channel receive-only head coil. NM-MRI data was acquired using a 2D magnetization transfer (MT) prepared gradient echo (GRE) sequence [13, 14]: echo time (TE) / repetition time (TR) = 2.68 ms / 337 ms, slice thickness 3 mm, in plane resolution 0.39×0.39 mm^2^, field of view (FOV) = 162 mm × 200 mm, flip angle (FA) = 40°, 470 Hz/pixel bandwidth, 15 contiguous slices, and magnetization transfer preparation pulse (300°, 1.2 kHz off resonance, 10 ms duration), 7 measurements, scan time 16 minutes 17 seconds. For registration from subject space to common space, a T_1_ magnetization-prepared rapid gradient echo (MP-RAGE) sequence was acquired with the following parameters: TE/TR= 3.02 ms/2600 ms, inversion time = 800 ms, FA=8°, voxel size = 1.0 × 1.0 × 1.0 mm^3^.

Cohort 2 was scanned with a Siemens Prisma 3 Tesla scanner using a 64-channel receive-only coil. NM-MRI data were acquired using a 2D GRE sequence with a MT preparation pulse: TE/TR = 3.10 ms /354 ms, 15 contiguous slices, FOV = 162×200 mm^2^, in plane resolution = 0.39 × 0.39 mm^2^), slice thickness = 3mm, 7 measurements, FA = 40°, 470 Hz/pixel receiver bandwidth, and MT pulse (300°, 1.2 kHz off resonance, 10 ms duration), scan time 17 minutes 12 seconds. For registration, structural images were acquired with an MP-RAGE sequence: TE/TR = 2.46 ms/1900 ms, inversion time = 900 ms, FA = 9°, voxel size = 0.8 × 0.8 × 0.8 mm^3^.

On the sagittal T_1_ images for both cohorts, the NM-MRI scan slices were positioned perpendicular to the dorsal edge of the brainstem at midline along the fourth ventricle, starting from the lower pons (below the most caudal extent of LC), with slices extending through the upper midbrain to cover both SNc and LC.

### Image Processing

MRI data was processed using the FMRIB Software Library (FSL). A transformation was derived between each individual’s T_1_-weighted image and 2 mm Montreal Neurological Institute (MNI) T_1_-space using FMRIB’s Linear Image Registration Tool (FLIRT) and FMRIB’s Nonlinear Image Registration Tool (FNIRT) in the FSL software package [41, 42]. Each participant’s T_1_-weighted image was brain extracted using the brain extraction tool (BET) in FSL. Next, an affine transform was used to align each participant’s brain extracted T_1_-weighted images with the MNI brain extracted image. Finally, a nonlinear transformation was used to generate a transformation from each participant’s T_1_-weighted images to T_1_-weighted MNI T_1_-space.

For each participant, individual NM-MRI measurements were corrected for motion by registering the seven measurements to the first image using a rigid-body transform in FLIRT and then averaged. Next, a transform was derived between each individual’s T_1_-weighted image and the averaged NM-MRI image with a boundary-based registration cost function.

SNc and LC volumes were segmented in native space using an automated thresholding method. To ensure consistent placement of reference regions of interest (ROIs), a reference ROI in the cerebral peduncle was created using the MNI template and the location of these reference ROIs are shown in blue in Fig 1. For each subject, the cerebral peduncle ROI was transformed to individual NM-MRI images using the MNI-T_1_ and T_1_-NM transforms described in previous paragraphs. The transform was done in a single step to reduce interpolation. The use of standard space ROIs ensured that the reference ROI was placed in similar locations for each subject. The mean (denoted μ_ref_), and standard deviation (σ_ref_) of the signal intensities were measured in the reference ROI. Next, standard space SNc and LC atlases, shown in Fig 1, were used to localize regions surrounding SNc and LC for thresholding [43]. These atlases were transformed from standard space to individual NM-MRI images, thresholded at a level of 5%, and dilated. The ROIs for thresholding were dilated to ensure that the entire SNc or LC were included for thresholding. The resulting ROIs in native space are shown in red in the middle column of Fig 1. Voxels in the resulting ROIs with intensity >μ_ref_+3.9σ_ref_ and >μ_ref_+2.8σ_ref_ were considered to be part of LC and SNc, respectively. A higher threshold was chosen for LC to compensate for susceptibility effects from the 4^th^ ventricle.

**Fig 1.**
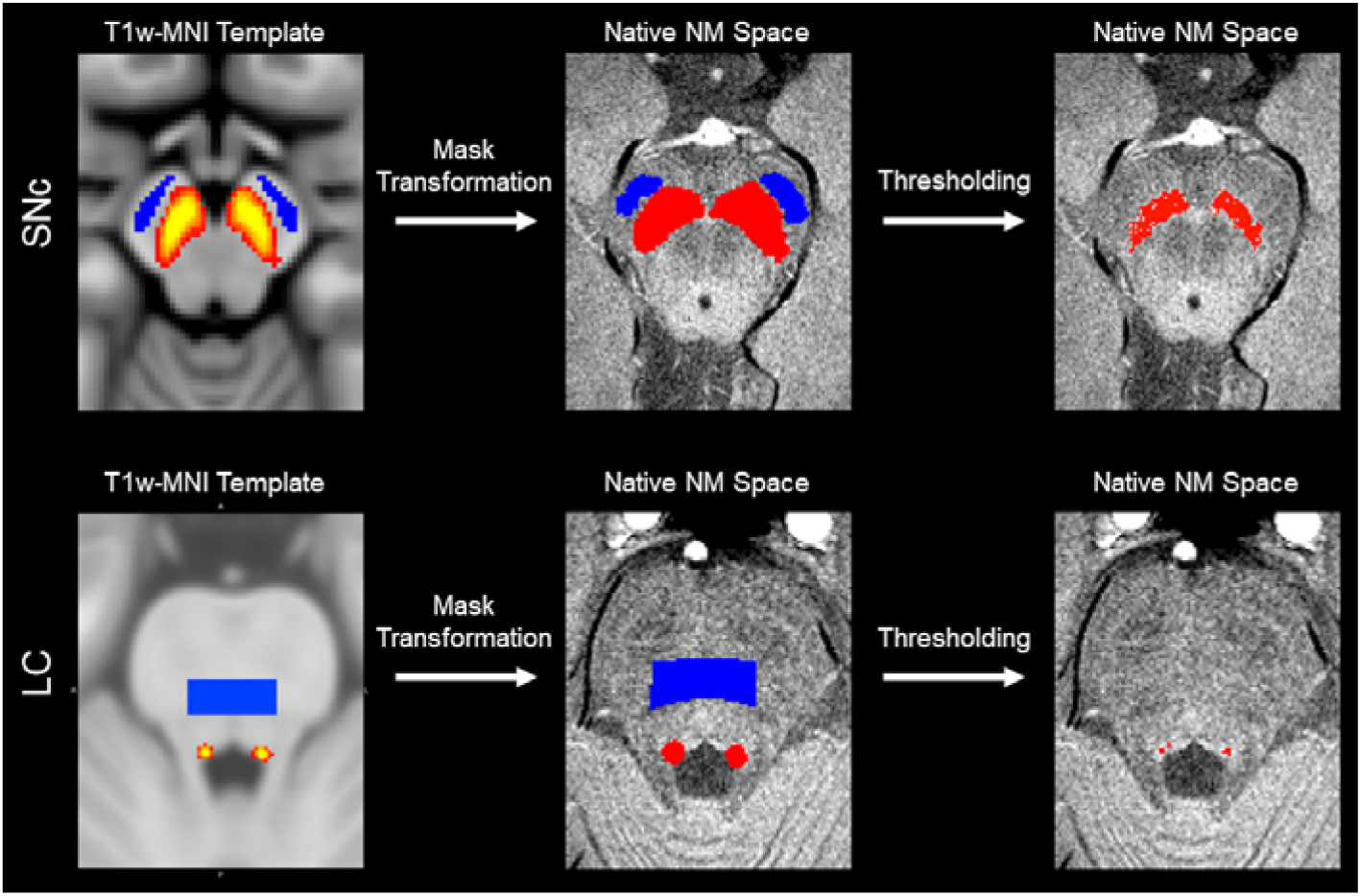
A pictorial representation of the segmentation procedure for SNc and LC. The first column shows the reference ROI in blue and the SNc atlas (top) or LC atlas (bottom) in red-yellow in MNI space. These ROIs were transformed to native space, and the atlases were threshold at a level of 5% and dilated (shown in red in the middle column). Voxels in the dilated region above the threshold were considered to be part of the SNc and LC (third column), respectively.

### Statistical Analysis

All statistical analyses were performed using IBM SPSS Statistics software version 24 (IBM Corporation, Somers, NY, USA) and results are reported as mean ± standard error. A *p* value of 0.05 was considered significant for all statistical tests performed in this work. Normality of SNc and LC volumes was assessed using the Shapiro-Wilk test for each group and all data was found to be normal.

For demographic data, independent samples t-test was used to assess differences in age and years of education and Chi square was used to examine differences in sex and race between controls and PD in each cohort. Group means for UPDRS-III score, disease duration, levodopa equivalents, MoCA, NMSQ, and RBD-SQ were compared using a two-tailed Welch’s *t*-test in each cohort.

In cohort 1, the effect of group (PD, control) was tested with separate analysis of covariance (ANCOVA) for SNc and LC volume controlling for age and education. In cohort 2, differences in SNc volume and LC volume between control and PD groups were compared using a two-tailed Welch’s t-test. Pearson’s correlation was used to assess the relationship between SNc volume with UPDRS-III, LEDD, MoCA and LC volume in PD for cohort 1, cohort 2, and both cohorts combined. Pearson’s correlation was used to assess the relationship between LC volume and 1) UPDRS-III, and 2) MoCA for cohort 1, cohort 2, and both cohorts combined. Age was a treated as a covariate in all correlations.

## Results

### Sample demographics

In cohort 1, significant differences were in age (*p*<10^−3^) and education (*p*=0.015) with controls being older and having higher levels of education than the PD group. No differences were seen between groups in sex (*p*=0.453) or in MoCA score (*p*=0.556). The PD group had significantly higher UPDRS-III (*p* <10^−3^), RBD-SQ (*p*=0.002), and NMSQ (*p*<10^−3^) scores compared to the control group. In cohort 2, no difference was seen between PD and control groups in age (*p*=0.894), race (*p*=0.402), sex (*p*=0.500), education (*p*=0.661), RBD-SQ score (*p*=0.087), or MoCA score (*p*=0.621). A significant difference was seen in NMSQ (*p*=0.0006) and UPDRS-III (*p*<10^−3^) scores between the two groups. Demographic information is summarized in **Table 1**.

**Table 1.** Demographic information for the groups used in this analysis. Data is presented as mean ± standard error unless noted otherwise. Two-tailed *t*-tests were used for group comparisons of age, education, MoCA, RBD-SQ, and NMSQ from which p values are shown. UPDRS-III was measured in the ON state. UPDRS-III - Unified Parkinson’s Disease Rating Scale Part III; MoCA – Montreal Cognitive Assessment; RBD-SQ - REM Sleep Behavior Disorder Screening Questionnaire; NMSQ - Non-motor Symptoms Questionnaire.

| Group characteristics | Cohort 1 |  |  | Cohort 2 |  |  |
| --- | --- | --- | --- | --- | --- | --- |
| | Control | PD | $P$ | Control | PD | $P$ |
| Participants | 19 | 22 | - | 33 | 39 | - |
| Age | 71.3 ± 1.2 | 60.4 ± 1.8 | $<10^{-3}$ | 63.5 ± 1.6 | 63.8 ± 1.6 | 0.894 |
| Sex (M:F) | 9:10 | 13:9 | 0.453 | 11:22 | 22:17 | 0.50 |
| Education [years] | 18.2 ± 0.5 | 16.3 ± 0.5 | **0.015** | 16.7 ± 0.4 | 17.0 ± 0.6 | 0.661 |
| Race [% Caucasian] | 78.9% | 100% | **0.023** | 90.6% | 87.2% | 0.402 |
| Disease Duration [years] | - | 6.8 ± 0.7 | - | - | 3.5 ± 0.6 | - |
| UPDRS-III | 3.2 ± 0.6 | 18.7 ± 2.3 | $<10^{-3}$ | 1.8 ± 1.9 | 18.9 ± 1.1 | $<10^{-3}$ |
| Levodopa Equivalents | - | 768.8 ± 90.4 | - | - | 623.3 ± 70.8 | - |
| MoCA | 27.5 ± 0.5 | 27.1 ± 0.6 | 0.566 | 28.0 ± 0.3 | 27.8 ± 0.3 | 0.621 |
| RBD-SQ | 2.3 ± 0.4 | 4.7 ± 0.6 | **0.002** | 2.7 ± 0.4 | 3.7 ± 0.4 | 0.087 |
| NMSQ | 3.4 ± 0.7 | 10.5 ± 0.9 | $<10^{-3}$ | 3.2 ± 0.5 | 6.6 ± 0.7 | **0.0006** |

### SNc and LC volume comparisons

Figs 2 and 3 show LC and SNc in mean MTC images of both cohorts. In cohort 1, the effect of group on volume was assessed with separate ANCOVAs for each ROI (SNc, LC) with age and education as covariates. In SNc, results revealed a significantly smaller volume in the PD group relative to the control group (Control: 474 mm^3^ ± 31 mm^3^; PD: 340 mm^3^ ± 28 mm^3^; *F*=8.031; *p*=0.007). Similarly, a reduction in LC volume was seen in the PD group relative to the control group (Control: 6.9 mm^3^ ± 0.7 mm^3^; PD: 4.4 mm^3^ ± 0.7 mm^3^; *F*=3.247; *p*=0.049). Age was not a significant variable in the ANCOVA model for LC volume (*p=*0.238) or SNc volume (*p=*0.08). Removal of non-Caucasian subjects did not change results, and reductions in LC volume (*p*=0.048) and SNc volume (*p*=0.005) were seen in the PD group relative to controls. In cohort 2, Welch’s *t*-test was used to examine group differences in SNc volume and LC volume. SNc (Control: 429 mm^3^ ± 20 mm^3^; PD: 329 mm^3^ ± 17 mm^3^; *t=*3.370; *p*=0.0002) and LC (Control: 8.0 mm^3^ ± 0.6 mm^3^; PD: 5.2 mm^3^ ± 0.6 mm^3^; *t=*3.306; *p*=0.0008) volumes were significantly lower in the PD group compared to the control group in cohort 2. These comparisons are shown in Fig 4.

**Fig 2.**
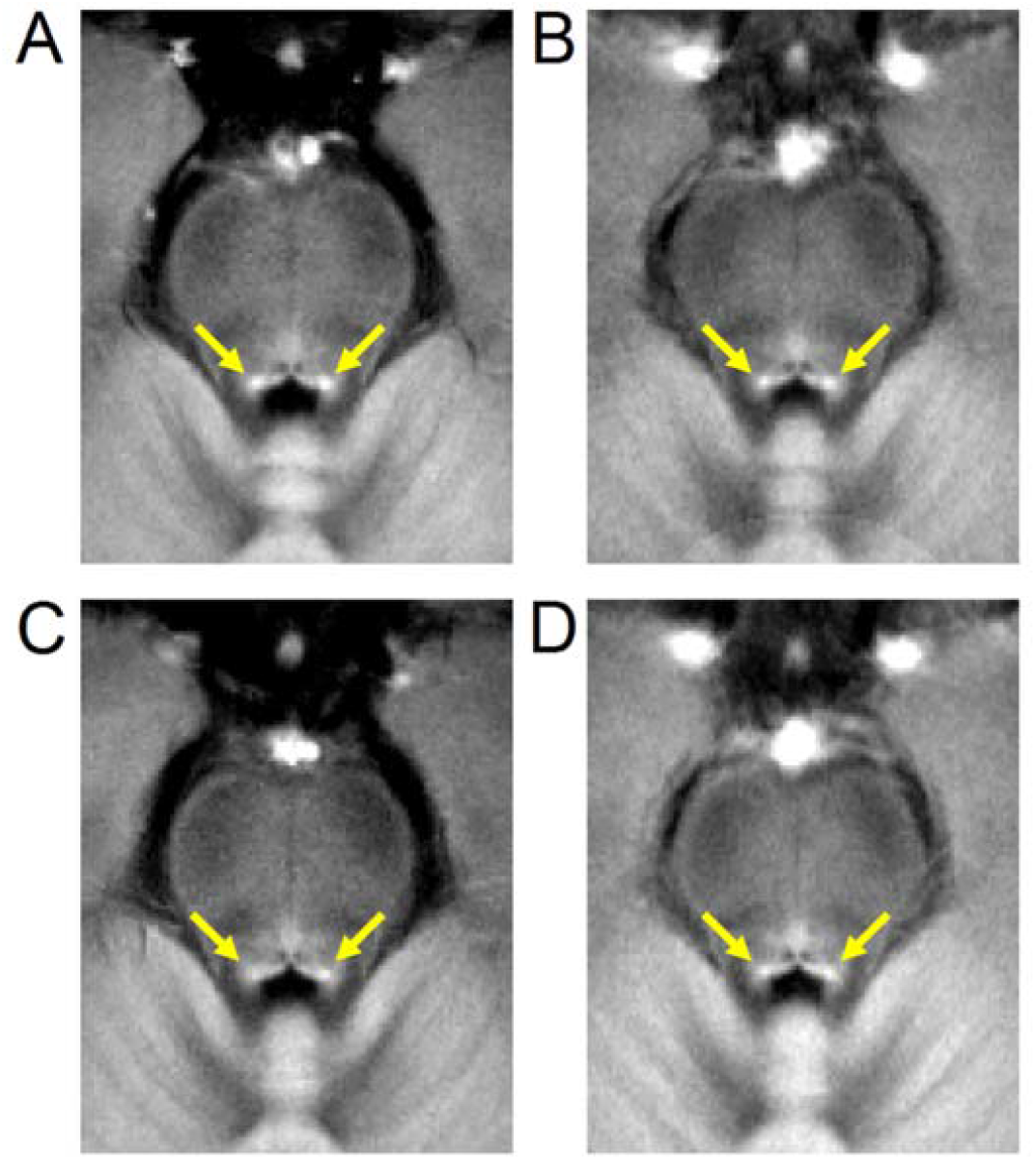
A comparison of mean LC contrast in control (top row) and PD (bottom row) groups for both cohorts. Mean MTC images from cohort 1 are shown in the left column while mean MTC images from cohort 2 are shown in the right column. For each group, the mean MTC image wa created by transforming MTC images from individual participants to MNI space and then averaging. In each image, yellow arrows indicate the location of LC.

**Fig 3.**
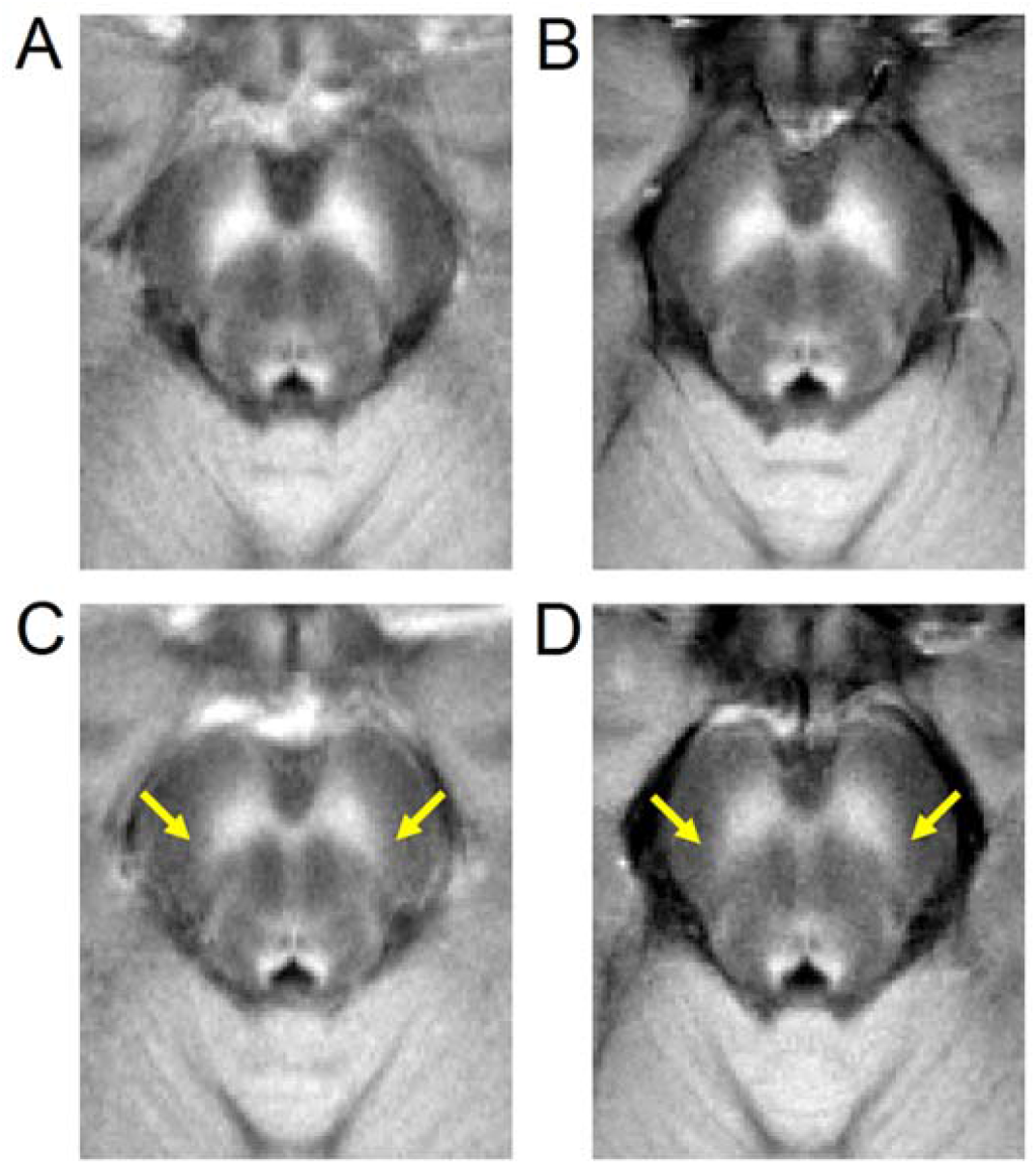
A comparison of mean SNc contrast in control (top row) and PD (bottom row) groups for both cohorts. Mean MTC images from cohort 1 are shown in the left column while mean MTC images from cohort 2 are shown in the right column. For each group, the mean MTC image wa created by transforming MTC images from individual participants to MNI space and then averaging. Regions experiencing PD-related neuronal loss are indicated by yellow arrows in the bottom row.

**Fig 4.**
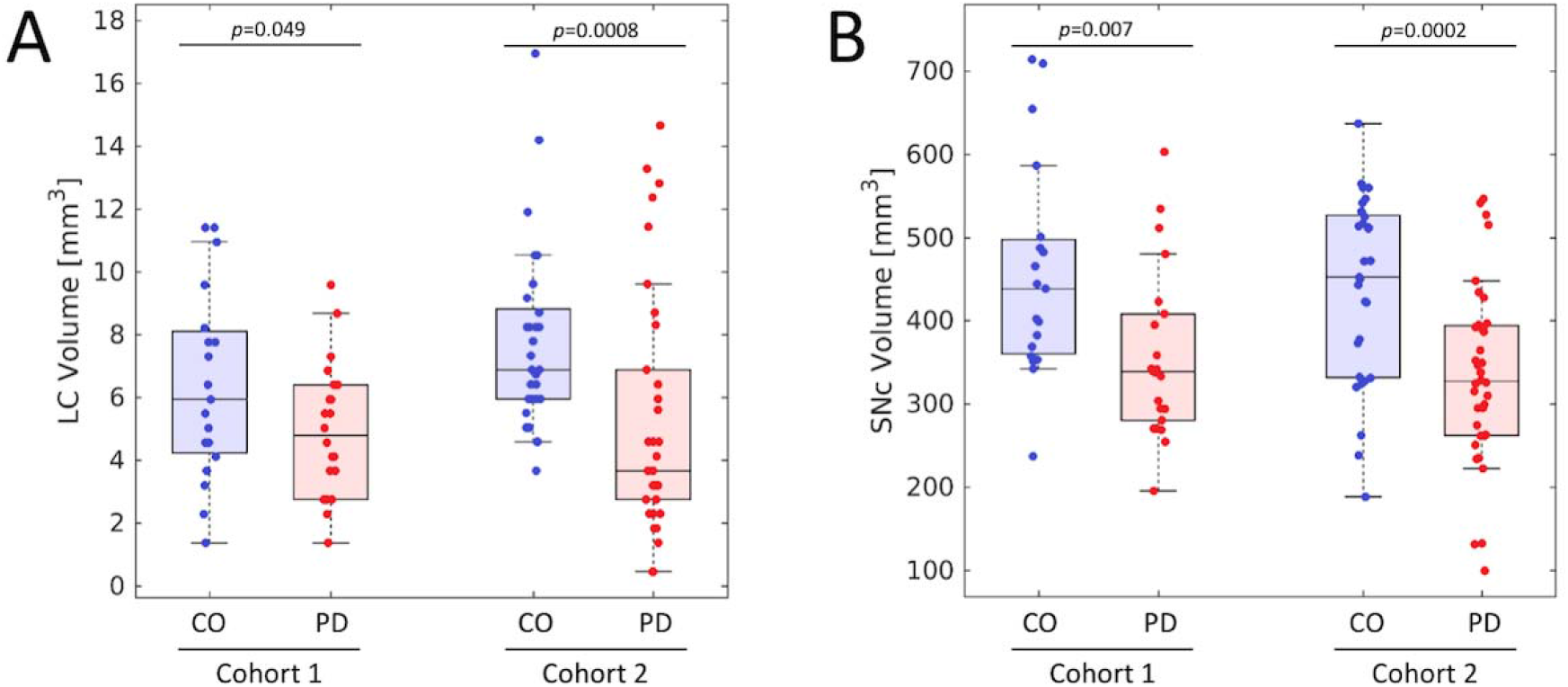
Group comparisons for LC volume (A) and SNc volume (B) in both cohorts. Similar reductions in LC and SNc volume are observed in both cohorts. In (A) and (B), the box denotes the 25^th^ and 75^th^ percentile, respectively, with the line denoting the median value.

### Clinical Correlations

In the PD group, MoCA showed a significant positive association with SNc volume in cohort 1 (*p*=0.012, *r*=0.478) and both cohorts (*p*=0.040, *r*=0.228) but not in cohort 2 (*p*=0.319, *r*=0.079). SNc volume showed no significant associations with UPDRS-III or disease duration in the PD group in cohort 1, cohort 2 or both cohorts (*ps*>0.436). There were no significant correlations between MoCA, UPDRS-III and disease duration with LC volume in cohort 1, cohort 2, or both cohorts combined for the PD group (*ps*>0.272). SNc volume showed a significant positive association with LC volume in PD in both cohorts (*p*=0.012, *r*=0.296) but no significance in HC (*p*=0.434, *r*=-0.025). In the PD group, SNc and LC volumes had a significant positive correlation in cohort 2 (*p*=0.024, *r*=0.333) but not in cohort 1 (*p*=0.138, *r*=0.243).

### ROC analysis

In cohort 1 SNc volume outperformed LC volume as a diagnostic imaging marker of PD. The area under the receiver operating characteristic (ROC) curve (AUC) for SNc volume was 0.756 [standard error (SE): 0.078, 95% confidence interval (CI): 0.603-0.909, *p*=0.005] while the AUC for LC volume was 0.644 [SE: 0.088, 95% CI: 0.471-0.816, *p*=0.117]. For cohort 1, combining SNc volume and LC volume yielded an AUC of 0.763 [SE: 0.074, 95% CI: 0.617-0.090, *p*=0.004]. NM-MRI measures of SNc volume and LC volume performed similarly in cohort 2 to differentiate PD from controls. LC volume had an AUC of 0.752 [SE: 0.063, 95% CI: 0.629-0.876, *p*=0.001] and SNc volume had an AUC of 0.749 [SE: 0.062, 95% CI: 0.627-0.871, *p*=0.001]. For cohort 2, combining SNc volume and LC volume yielded an AUC of 0.775 [SE: 0.057, 95% CI: 0.664-0.886]. ROC curves for both cohorts are shown in Fig 5.

**Fig 5.**
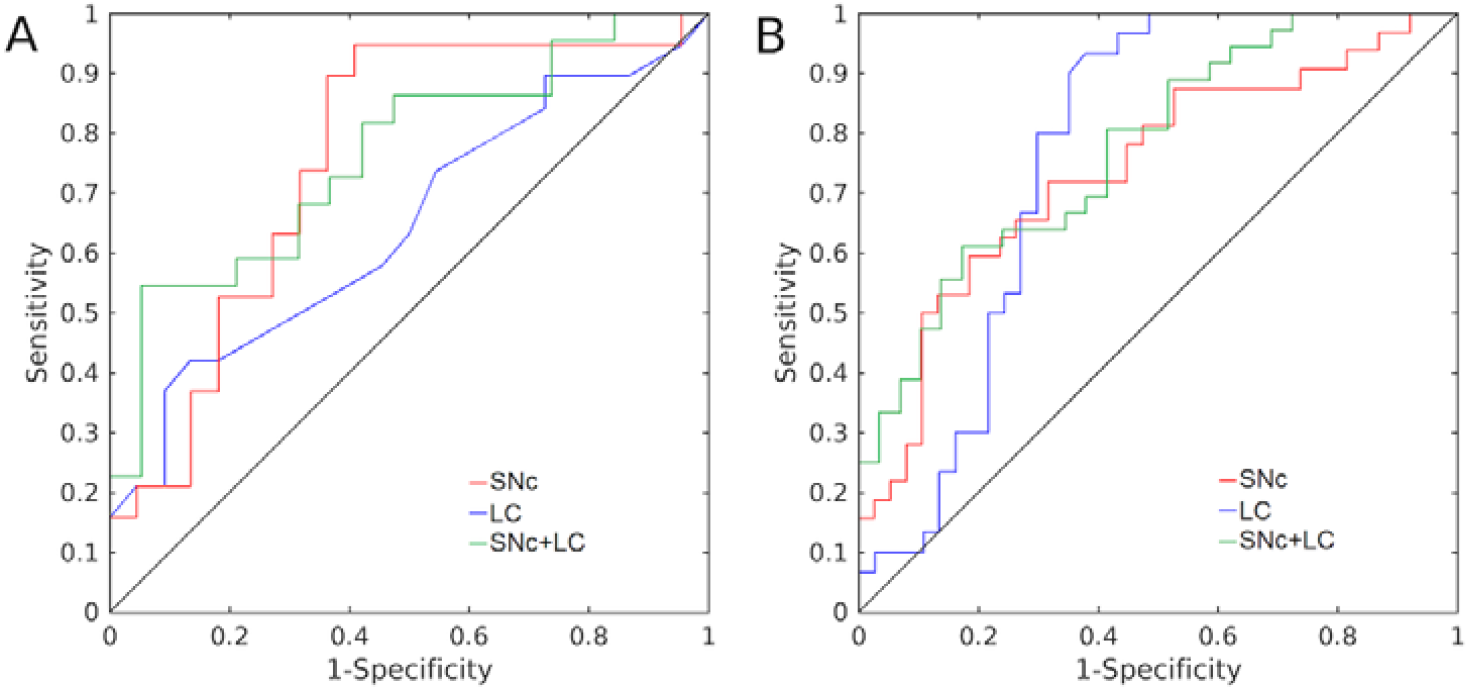
ROC analysis assessing performance of SNc and LC volumes to differentiate PD from controls. ROC curves for Cohort 1 and Cohort 2 are shown in (A) and (B), respectively.

## Discussion

This study examined PD-related loss of NM-MRI contrast in LC and SNc in two separate cohorts, using two different scanner models. We observed significant volume loss in both LC and SNc in the PD groups as compared to controls in both cohorts. In cohort 2, SNc volume and LC volume performed similarly as diagnostic imaging markers of PD. This is the first report, to our knowledge, of reproducible detection of PD-associated LC volume loss using the same NM-MRI approach in multiple cohorts. It is also the first report of simultaneous imaging of LC and SNc detecting PD effects in both structures in discovery and validation cohorts. The NM-MRI pulse sequence and image processing methods that were used in this study have previously established high scan-rescan reproducibility [18, 20]. To increase reproducibility and ensure consistent placement of ROIs, reference regions and regions within which to threshold voxels for volume measurement were defined using standard space ROIs and transformed to each individual’s NM-MRI image. These ROIs were then used to define thresholded and reference regions in the SNc and LC segmentation procedure. The results of this study, therefore, support the utility of these NM-MRI methods as a robust and reproducible approach to measure PD neuropathology *in vivo*.

The two Parkinsonian cohorts used in this study had similar motor impairment, but different disease durations. The mean and standard error of nigral volume was strikingly consistent across both cohorts. The consistency in SNc volume loss may be due to similarity of NM-MRI protocols used in this study. A previous study assessed LC volume measurements using multiple types of scanners [34], but used different implementations of NM-MRI protocols to image LC. The authors found differences in NM-MRI measures of LC between scanners, but it was not clear whether these differences resulted from the variability in scanning protocols or the scanners themselves. Here, we observed a non-significant trend toward an LC volume difference between the two cohorts in the control groups (*p*=0.081). Because the NM-MRI protocols were very similar, this result suggests that differences between the Prisma and Trio scanner types (the former having higher signal to noise ratio due to signal digitization at the receiver coil) may enable the Prisma to detect NM-MRI contrast in LC more readily than the Trio scanner. This result should be interpreted cautiously though, because an alternative explanation is that the older control group in cohort 1 may have had lower LC volume than cohort 2 due to aging effects.

The performance of cohort 1 LC volume as a diagnostic marker of PD was similar to the AUC reported in a previous study, and cohort 2 LC volume outperformed that observed in the prior study [34]. Improved performance of LC volume in cohort 2 may be due to several factors. First, the Siemens Trio MRI scanner used in cohort 1 has a higher noise profile than the Siemens Prisma MRI scanner used in cohort 2, and elevated noise likely reduced efficacy of LC volume as a diagnostic marker in cohort 1. Second the control group was significantly older than the PD group in cohort 1. Neuromelanin peaks in LC between age 50-60 and declines after age 60 [44-46]. Thus, this age difference may have reduced the effect size by reducing the mean LC volume in the control group.

The ROC analysis found AUC of nigral volume in both cohorts to have AUCs comparable to previously published nigral diagnostic imaging markers of PD [27, 34, 47-50]. However, these AUCs are below thresholds typically used for individual clinical diagnosis (AUC > 0.9). Combining LC and nigral volume increased the AUC in both cohorts (AUC = 0.763 and AUC = 0.775 for cohorts 1 and 2, respectively) and these values are approaching levels that are clinically useful. Diagnostic accuracy may be improved by combining nigral markers from other MRI contrasts, such as those from iron [28, 31, 32, 51] or diffusion [29, 52] contrasts.

Nigrosome-1 is the subregion of SNc that experiences the greatest reductions in melanized neurons and is found in the lateral-ventral portion of SNc [53, 54]. This region can be visualized in T_2_- or T_2_*-weighted images as a relatively hyperintense region in the substantia nigra, and loss of this hyperintensity in PD patients has shown promise as a diagnostic imaging marker [55]. In NM-MRI images, nigrosome-1 has been localized to the lateral-ventral portion of SNc, which is consistent with its localization in postmortem neuropathology studies [3, 53, 56]. We observed reductions in NM-sensitive contrast in the lateral-ventral regions of SNc (see Fig 3) in both cohorts. This is in agreement with earlier studies that found volume loss or reductions in neuromelanin-sensitive contrast in the posterior portion of SNc [57] or in the lateral-ventral tier of SNc [27]. Studies examining PD-related changes using other MRI contrasts have found increases in free water [58, 59], which may be associated with neurodegeneration [60], in posterior nigral ROIs as well as elevated iron levels in the lateral-ventral tier [61, 62]. Taken together, these changes may manifest from neurodegeneration in nigrosome-1.

There are several caveats to this study. First, UPDRS-III was measured in the on-medication state. Earlier studies found a significant relationship between nigral volume and UPDRS score [34] and lack of correlation between nigral volume and UPDRS-III score may be due to measurement in the on-medication state. This may also have been due to the inclusion of PD patients with similar early to moderate disease stage, and a lack of PD patients with more severe motor symptoms. Second, the control group in cohort 1 was significantly older and more educated than the PD group. This may have reduced the effect size for LC volume. Finally, we elected to use 3 mm slices in the acquisition to ensure sufficient signal to noise ratio (SNR) for SNc and LC segmentation, which may increase partial volume effects.

NM-MRI approaches based on explicit magnetization transfer effects have already demonstrated high scan-rescan reproducibility [18-20]. The current findings provide additional evidence that NM-MRI robustly and reproducibly detects PD-related reductions in volume in both LC and SNc. In the context of emerging evidence for a mechanistic role for neuromelanin in PD [8-10], these findings may inform strategies to study disease pathophysiology and ultimately assist therapeutics development. Therefore, investigation of these NM-MRI measures as candidate biomarkers in larger, longitudinal studies is warranted. They have potential for development both as individual markers and as part of multivariate profiles incorporating iron-sensitive and diffusion-sensitive MRI modalities.

## Data Availability

All data produced in the present study are available upon reasonable request to the authors

## Author Roles

1) Research project: A. Conception, B. Organization, C. Execution; 2) Statistical Analysis: A. Design, B. Execution, C. Review and Critique; 3) Manuscript: A. Writing of the first draft, B. Review and Critique.

K.H. 1C, 2A, 2B, 2C, 3A, 3B

J.L.: 1A, 1B, 1C, 2A, 2C, 2B, 3A, 3B R.T.: 2B, 2C, 3B

X.H: 1A, 1C, 2C, 3B

D.H.: 1A, 1B, 1C, 2A, 2B, 2C, 3A, 3B

## Acknowledgements

The authors would like to express their gratitude to Ms. Kelsey Tucker for assistance with data acquisition.

